# CMANet: Cross-Modal Attention Network for 3-D Knee MRI and Report-Guided Osteoarthritis Assessment

**DOI:** 10.1101/2025.06.08.25329119

**Authors:** R. Harari, H. Rajabzadeh-Oghaz, F. Hosseini, M. Gholami Shali, A. Altaweel, N. Haouchine, F. Rikhtegar

## Abstract

**Objective:** Knee osteoarthritis (OA) is a leading cause of disability worldwide, with early identification of structural changes critical for improving patient outcomes. While magnetic resonance imaging (MRI) provides rich spatial detail, its interpretation remains challenging due to complex anatomy, subtle lesion presentation, and limited voxel-level annotations. Meanwhile, radiology reports encode semantic and diagnostic insights that are typically underutilized in imaging AI pipelines. In this work, we introduce CMANet, a Cross-Modal Attention Network that integrates 3D knee MRI volumes with their corresponding free-text radiology reports for joint OA severity classification and lesion segmentation. CMANet introduces four key innovations: (1) an asymmetric cross-modal attention mechanism that enables bidirectional information flow between image and text, (2) a weakly supervised anatomical alignment module linking report phrases to MRI regions, (3) a multi-task prediction head for simultaneous OA grading and voxel-level lesion detection, and (4) interpretable attention pathways for tracing predictions to report language and anatomical structures. Evaluated on a dataset of 642 patients with paired MRI and radiology reports, CMANet achieved significant improvements over unimodal baselines—boosting KL-grade classification AUC from 0.769 to 0.871 (Δ =0.102, p=0.004) and increasing Dice scores for cartilage and BML lesion segmentation. The model also demonstrated generalizability in predicting 2-year OA progression (AUC=0.804) and achieved improved alignment between anatomical regions and textual descriptions. These results highlight the potential of multimodal learning to enhance diagnostic accuracy, spatial localization, and explainability in musculoskeletal imaging.

## 1 Introduction

Medical imaging plays a critical role in the diagnosis, prognosis, and monitoring of musculoskeletal diseases. Among these, osteoarthritis (OA) remains a major global health burden, affecting over 300 million individuals and leading to chronic pain, reduced mobility, and diminished quality of life [1,2]. The knee is the most commonly affected joint, and early identification of disease progression is essential for informing therapeutic decisions and improving long-term outcomes [3].

Magnetic resonance imaging (MRI) is widely regarded the most sensitive modality to visualize early structural changes in OA [4], including cartilage thinning [5,6], bone marrow lesions (BML) [7], meniscal tears [8], and synoviti [9], long before such changes manifest on radiographs. However, the high-dimensional, multi-planar nature of MRI data and the subtlety of early-stage lesions pose significant challenges for automated analysis [10]. Furthermore, expert annotations for voxel-level segmentation are time-consuming and often unavailable at scale [4].

In parallel, clinical workflows routinely generate radiology reports that summarize key pathological findings using structured medical language [11]. These free-text reports capture semantic cues, diagnostic impressions, and anatomical references that may not be explicitly labeled in the imaging data [12]. Despite their richness, these reports are underutilized in most imaging AI pipelines, which typically rely on either purely image-based methods or isolated natural language processing (NLP) models. The lack of integrated frameworks that jointly reason over image and text modalities represents a missed opportunity for improving model performance, interpretability, and clinical alignment.

Recent work in multimodal learning has shown the potential for aligning vision and language representations in natural and biomedical domains [13]. However, existing approaches are often limited by one or more of the following: (i) reliance on dense supervision or manual region-phrase annotations [14], (ii) lack of spatial grounding in attention mechanisms [15], and (iii) symmetric fusion schemes that treat modalities equally despite inherent differences in dimensionality and informativeness [16]. These limitations are especially salient in musculoskeletal applications, where pathology is localized, language is often sparse, and anatomy is complex.

To address these gaps, we introduce **CMANet**—a Cross-Modal Attention Network that integrates 3D MRI volumes and their corresponding radiology reports for OA severity grading and lesion segmentation. CMANet incorporates four key innovations: (1) an asymmetric cross-modal attention mechanism that uses report context to guide spatial focus in MRI and vice versa; (2) an anatomical alignment module that leverages weak region-phrase supervision using automated segmentation and entity recognition; (3) a multi-task learning architecture optimized for both global (classification) and local (segmentation) tasks; and (4) interpretability mechanisms that enable token-to-region and region-to-phrase tracing.

We evaluate CMANet on a large cohort of 642 knee OA patients with paired MRI and radiology reports, including a subset with lesion-level annotations.

## 2 Method

We propose a CMANet designed to integrate volumetric MRI scans and free-text radiology reports for the joint tasks of OA severity classification and structural lesion segmentation. The architecture addresses key challenges in clinical imaging: weak supervision, modality heterogeneity, and sparse regional annotations. CMANet unifies volumetric visual encoding, biomedical language modeling, cross-modal attention, anatomical alignment, and multi-task prediction in an end-to-end trainable framework.

Let *V* ∈ R^*D*×*H*×*W*×*C*^ denote the input 3D MRI volume, where *D* is the number of slices, *H* and *W* are spatial dimensions, and *C* is the number of channels (T2-weighted sagittal and coronal sequences). Let *R* = {*w*_1_,*w*_2_,…,*w*_*n*_} represent the associated radiology report, tokenized into *n* subword units. CMANet outputs (1) a scalar classification prediction *ŷ*_cls_ ^∈^ R^*C*^ over OA grades (KL 0–4), and (2) a 3D segmentation mask *ŷ*_seg_ ∈ [0,1]^*D*′×*H*′×*W*′^ highlighting lesion locations such as cartilage damage or BMLs.

The model (Figure1) illustrates the ROC curves. comprises five core components: (i) a 3D CNN-based MRI encoder, (ii) a transformer-based report encoder, (iii) a bidirectional cross-modal attention module, (iv) a phrase-region anatomical alignment module, and (v) a multi-task output head for classification and segmentation.

**Figure 1.**
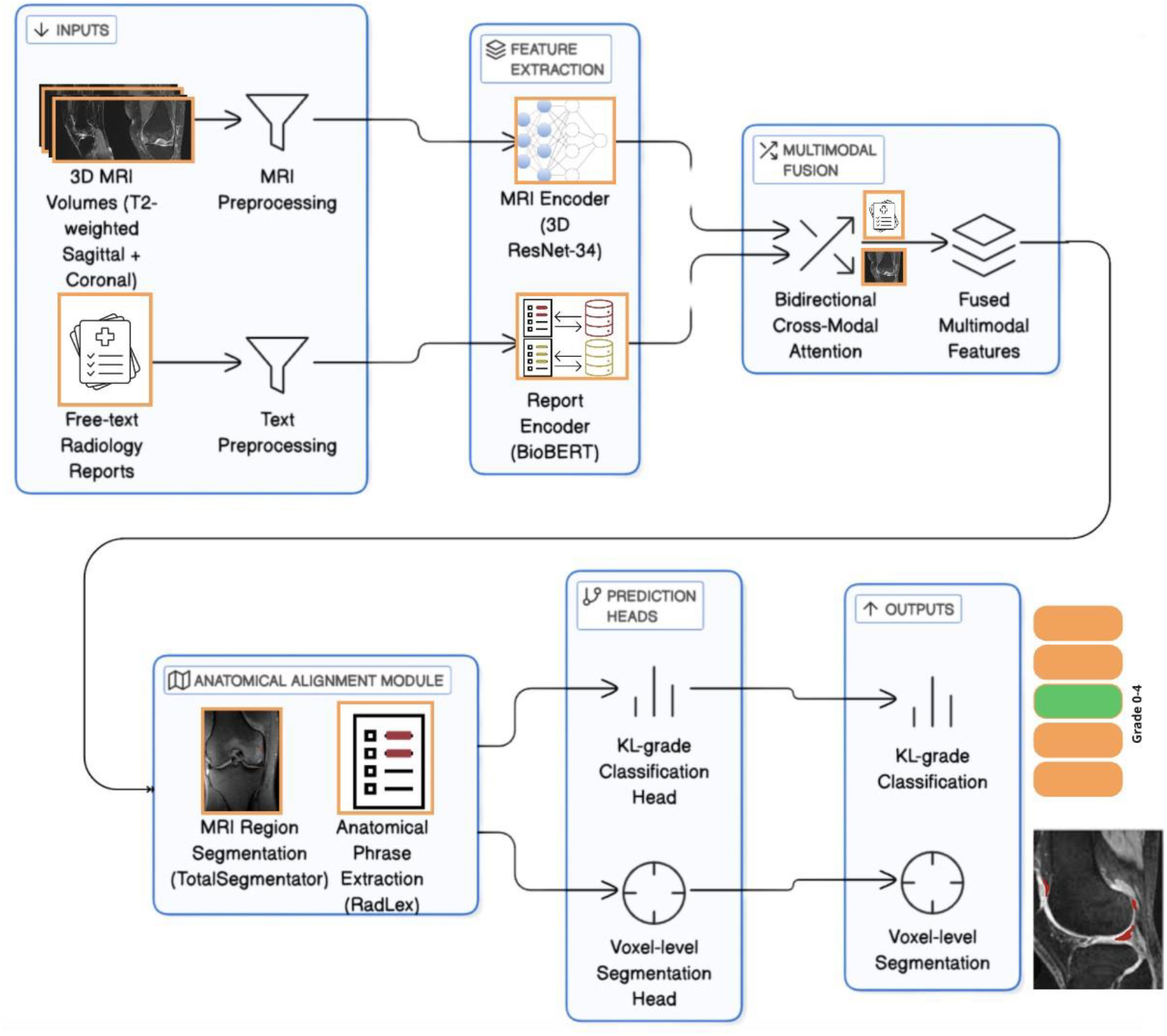
CMANet architecture for knee OA assessment. 3D MRI and radiology reports are processed via a 3D ResNet-34 and BioBERT, then fused using cross-modal attention. An anatomical alignment module links report phrases to MRI regions. Outputs include KL-grade classification and voxel-level lesion segmentation.

### 2.1 Volumetric MRI Encoder

**T**he knee joint exhibits complex anatomical structures across multiple planes, and disease manifestations such as cartilage thinning, subchondral bone changes, and effusions are distributed non-locally. Capturing these patterns requires spatially coherent feature representations derived from the full MRI volume. We used a deep 3D convolutional model based on ResNet-34, adapted for medical imaging, to hierarchically encode these features.

**E**ach MRI volume undergoes standard corrections including N4 bias field correction, rigid alignment of sagittal and coronal sequences, and voxel resampling to an isotropic 1 mm^3^ grid. The sagittal and coronal T2-weighted sequences are co-registered and stacked as separate channels, creating an input tensor *V* ^∈^ R^*D*×*H*×*W*×2^.

#### Initial feature extraction

The network begins with a 3D convolutional layer followed by batch normalization, ReLU activation, and max pooling:

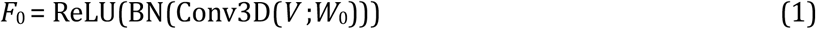

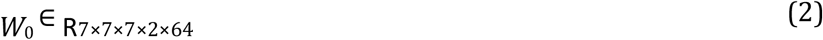

Max pooling with kernel size 3 × 3 × 3 and stride 2 is then applied to reduce the spatial resolution and increase receptive field.

#### Residual encoding

The processed tensor is passed through four residual stages, each comprising multiple convolutional blocks:

- Stage 1: 3 blocks, 64 channels
- Stage 2: 4 blocks, 128 channels
- Stage 3: 6 blocks, 256 channels
- Stage 4: 3 blocks, 512 channels

Each block includes two 3 × 3 × 3 convolutions with batch normalization and ReLU, and a skip connection that either passes the identity or applies a 1×1×1 convolution for downsampling when input and output dimensions differ.

#### Learned relevance masking

To encourage anatomical specificity, we introduce a side branch that learns a soft spatial mask *M*(*V*) ∈ [0,1]^*D*×*H*×*W*^. This mask is computed via a pair of 3D convolutions with sigmoid activation:

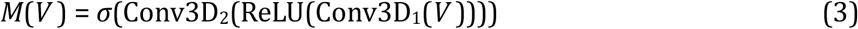

The input volume is then element-wise reweighted:

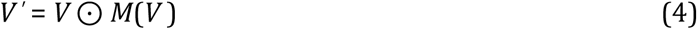

This masked volume *V* ^′^ is used as an input to the residual encoder, to enable suppression of background voxels and improving attention localization.

#### Output

The final output of the MRI encoder is a feature tensor *F*_MRI_ ∈ R^*D*′×*H*′×*W*′×512^, where *D*^′^, *H*^′^, and *W*^′^ depend on the downsampling factors of the network.

### 2.2 Radiology Report Encoder

**R**adiology reports provide structured yet sparse descriptions of pathology, often containing diagnostic conclusions, severity descriptors, and anatomical localization. These textual elements offer complementary supervision to imaging but require contextual embedding to resolve ambiguity. We utilize BioBERT, a pre-trained biomedical transformer, to convert each report into a highdimensional semantic representation. We further extract anatomical phrases from the report to align with imaging regions in a weakly supervised manner.

#### Preprocessing

Each report *R* is cleaned to remove boilerplate headers and institution-specific text, then tokenized using the WordPiece tokenizer employed by BioBERT. The resulting token sequence is *R* = {*w*_1_,*w*_2_,…,*w*_*n*_} where *n* varies by report. The tokens are embedded using a frozen BioBERT model:

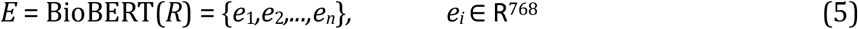

#### Global embedding

To produce a fixed-length representation of the entire report, we apply global average pooling:

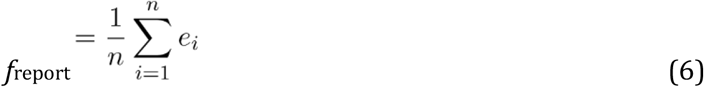

This vector captures a holistic view of the radiologist’s findings, and is used to condition the crossmodal attention mechanism (see Sec. 2.3).

#### Anatomical phrase embeddings

To enable anatomical grounding, we identify *K* meaningful phrases *p*_*k*_ in the report corresponding to regions or pathologies (e.g., “medial femoral condyle,” “subchondral edema”). This is accomplished using a rule-based entity recognition pipeline built on RadLex and SNOMED-CT terminologies.

Each phrase is defined by its token span (*i*_*k*_,*j*_*k*_) and embedded by averaging its constituent token embeddings:

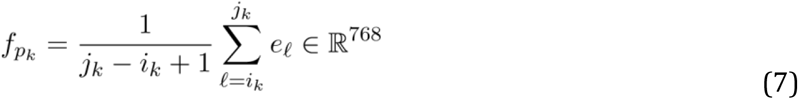

These phrase vectors are used later in the anatomical alignment module (Sec. 2.4) to provide region-level supervision without explicit annotations.

#### Output

The report encoder creates a global vector *f*_report_ and a set of anatomical phrase embeddings 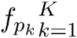. Both are passed to downstream attention and alignment modules.

### 2.3 Asymmetric Cross-Modal Attention

**T**he MRI and report modalities exhibit structural asymmetry: the former is spatially dense and high-dimensional, while the latter is sequential, sparse, and information-rich. Instead of fusing modalities symmetrically, we adopt an asymmetric attention mechanism that (1) allows the global report context to guide spatial focus over MRI, and (2) enables the MRI content to re-weight textual features based on visual salience. This bidirectional attention enhances interpretability and improves multimodal grounding.

#### Report-to-MRI attention

The global report embedding *f*_report_ ∈ R^*d*^ is first projected into the query space:

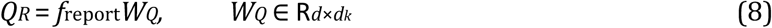

MRI features *F*_MRI_ ∈ R^*D*′×*H*′×*W*′×*C*′^ are projected voxel-wise to keys and values:

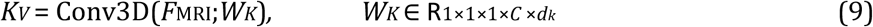

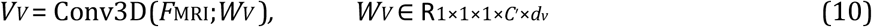

At each spatial location (*x,y,z*), we compute the attention weight:

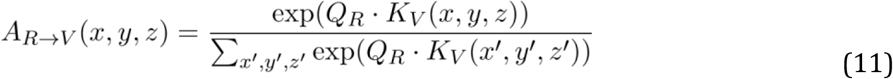

The output is an attended MRI representation:

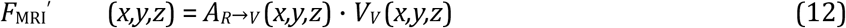

#### MRI-to-report attention

We extract global visual context by applying global average pooling to *F*_MRI_ and project it into a query vector:

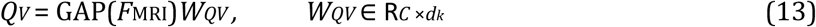

The full report embeddings *E* = {*e*_1_,…,*e*_*n*_} are projected into key and value spaces:

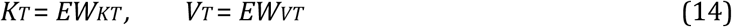

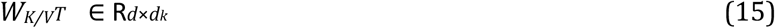

Attention over report tokens is computed as:

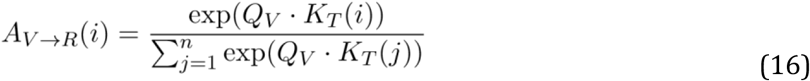

The resulting attention-weighted report vector is:

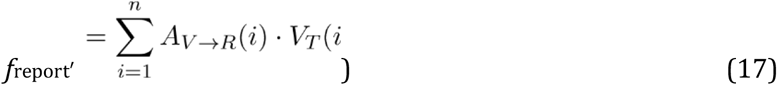

#### Output

The result of this module is a refined MRI feature map *F*_MRI_^′^ and an updated global report vector *f*_report_^′^, each modulated by the complementary modality.

### 2.4 Anatomical Alignment via Weak Phrase-to-Region Supervision

**R**adiology reports often mention anatomical structures and pathologies (e.g., “cartilage thinning in the medial femoral condyle”) without spatial annotations. To leverage these signals for regionspecific supervision, we introduce a weak alignment mechanism that associates textual phrases with segmented anatomical regions in the MRI. This phrase-to-region supervision constrains the learned representations to preserve spatial interpretability.

#### Region segmentation

We leverage the TotalSegmentator framework to obtain anatomical segmentations 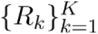 from the MRI volume, corresponding to distinct knee compartments and structures (e.g., patella, medial tibial plateau). Each *R*_*k*_ is a binary mask indicating the voxels of the *k*-th anatomical region.

#### Region-level feature pooling

For each anatomical region *R*_*k*_, we compute a mean feature representation from the encoded MRI feature map *F*_MRI_:

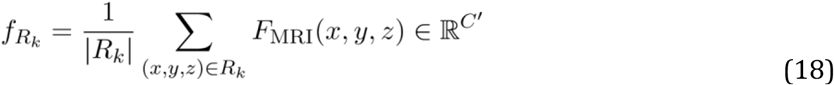

#### Phrase-region alignment loss

From the report, we extract anatomical phrases {*p*_*k*_} and their embeddings 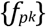 as described in Sec. 2.2. Each phrase is assumed to correspond to one region. We define a cosine alignment loss between the MRI-derived and text-derived representations:

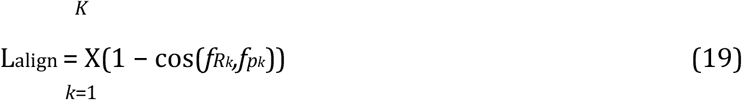

This weakly encourages the model to co-embed spatial and semantic information, improving anatomical localization without explicit voxel-level text supervision.

#### Implementation note

For each sample, we restrict alignment to only regions and phrases that co-occur. For ambiguous or overlapping phrases (e.g., “joint space”), we allow soft many-tomany alignment by averaging matched regions or phrases.

#### Output

This alignment term augments the model’s training objective and reinforces anatomical interpretability of both the attention and segmentation outputs.

### 2.5 Multi-Task Prediction Heads

**O**A evaluation includes two main tasks: global grading of disease severity (e.g., using the Kellgren– Lawrence scale) and spatially localized identification of structural lesions such as bone marrow lesions (BMLs), cartilage defects, and synovitis. To support clinical workflows, CMANet is designed with dual prediction heads: a classification branch and a segmentation branch, both trained from shared multimodal features.

#### Feature fusion

The attended MRI feature map 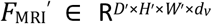 is globally pooled and concatenated with the attention-weighted report vector 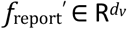 to form a fused descriptor:

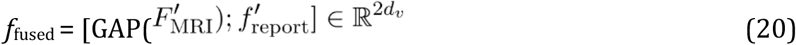

This vector aggregates spatial and semantic context for global classification.

#### Classification head

OA severity is predicted as a categorical distribution using a fully connected softmax layer:

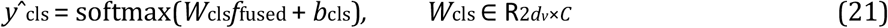

where *C* is the number of KL grades (5 for grades 0–4). We add dropout (*p* = 0.3) for regularization before the output layer.

#### Segmentation head

For voxel-level prediction, we apply a 1 × 1 × 1 convolution to the attended MRI feature map, followed by a sigmoid activation:

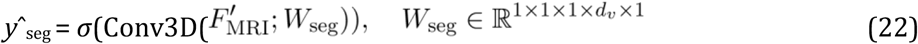

This produces a binary mask *ŷ*_seg_ ∈ [0,1]^*D*′×*H*′×*W*′^ indicating lesion presence probability at each voxel.

#### Interpretability hook

Both output heads can be traced back through the cross-modal attention weights (Sec. 2.3), enabling token-to-region visualization and region-to-phrase saliency analysis, useful for post hoc clinical review.

### 2.6 Loss Functions and Optimization

#### Composite learning objective

CMANet is trained end-to-end using a multi-task loss function that combines four components: classification loss for OA grading, segmentation loss for voxel-wise lesion detection, alignment loss for anatomical grounding, and an auxiliary attention regularization loss to promote focused cross-modal interactions.

#### Classification loss

For the OA severity task, we use the standard categorical cross-entropy loss:

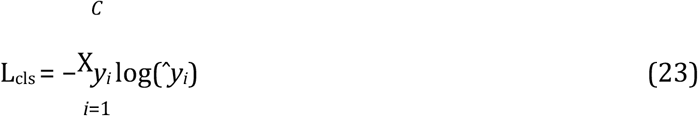

where *y* ∈ {0,1}^*C*^ is the one-hot ground truth vector, and *ŷ* ∈ [0,1]^*C*^ is the softmax output from the classification head.

#### Segmentation loss

For lesion segmentation, we use the Dice loss:

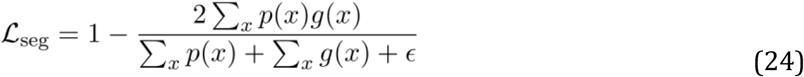

where *p*(*x*) = *ŷ*_seg_(*x*) is the predicted probability at voxel *x, g*(*x*) is the binary ground truth label, and *ϵ* is a small constant (e.g., 10^−5^) for numerical stability.

#### Alignment loss

The anatomical alignment loss L_align_ is defined in Sec. 2.4 as the average cosine distance between phrase embeddings and their matched region embeddings:

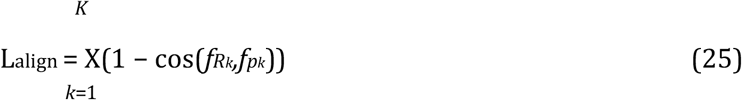

#### Attention sparsity regularization

To prevent attention maps from collapsing into uniform distributions or overfitting to noisy tokens, we add an entropy-based regularization term on the report-to-MRI attention weights:

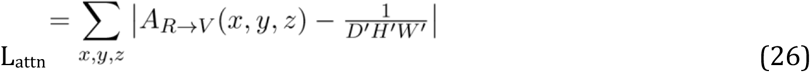

#### Total loss

The final training objective is a weighted sum of the above components:

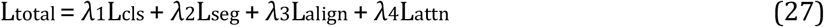

where *λ*_1_,*λ*_2_,*λ*_3_,*λ*_4_ are scalar weights (default values: 1.0,1.0,0.5,0.01 respectively), selected via grid search on the validation set.

#### Optimization

We optimize L_total_ using the Adam optimizer with learning rate 1 × 10^−3^, *β*_1_ = 0.9, *β*_2_ = 0.999, and weight decay 1 × 10^−5^. Training is performed for 100 epochs with early stopping (patience = 10) on the validation loss.

### 2.7 Implementation and Experimental Setup

#### Hardware and framework

All experiments were conducted on a cluster of four NVIDIA RTX A6000 GPUs (48 GB each). The model was implemented in PyTorch v2.0 with CUDA 11.8 and trained using mixed precision (AMP) to optimize memory usage. Transformer components were integrated from the HuggingFace Transformers library v4.35.

#### Preprocessing pipeline

MRI volumes were bias-corrected using the N4ITK algorithm, then resampled to an isotropic resolution of 1 mm^3^ using linear interpolation. T2-weighted sagittal and coronal sequences were rigidly co-registered using ANTs. Volumes were cropped or padded to a fixed size of *D* × *H* × *W* = 64 × 256 × 256 and intensity-normalized (z-scoring). Reports were cleaned to remove headers, tokenized using BioBERT’s WordPiece tokenizer, and truncated to 512 tokens if necessary.

#### Dataset and split

We curated a dataset of 642 patients with paired knee MRI scans and corresponding radiology reports. A patient-level split was used to prevent information leakage: 70% for training (450 patients), 15% for validation (96 patients), and 15% for testing (96 patients). A subset of 120 patients included longitudinal follow-ups (24 months intervals), used for downstream generalization studies.

#### Training protocol

We trained CMANet for 100 epochs with batch size 8 and early stopping based on validation loss. Dropout (*p* = 0.3) was applied to all fully connected layers and the attention module. All weights were initialized using He initialization. Learning rate was scheduled with cosine annealing and warm restarts every 20 epochs.

#### Inference and reproducibility

During inference, predictions were computed slice-wise and post-processed using connected-component filtering to eliminate spurious lesion clusters. All hyperparameters, code, trained checkpoints, and preprocessing scripts will be made publicly available via GitHub and HuggingFace Model Hub upon publication.

## 3 Results

### 3.1 KL-Grade Classification Results

We first compare CMANet with two unimodal baselines in the 96-patient held test cohort (Table1). CMANet increases macro AUC from 0.769 to 0.871 (Δ = 0.102, p = 0.004, DeLong test) and macro-F_1_ from 0.634 to 0.689. The improvement in macro-AUC is robust: 0.871 [0.81–0.92] vs 0.769 [0.70– 0.83]. Figure2 illustrates the ROC curves.

**Table 1:**
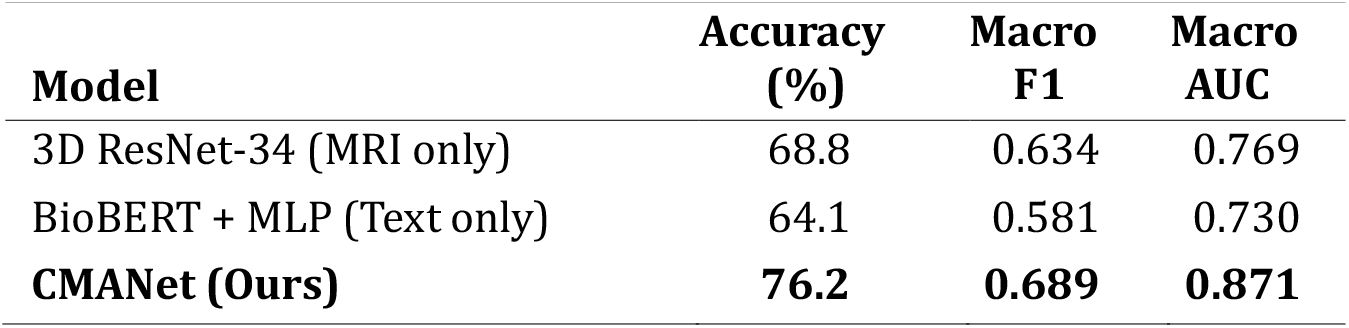
KL-Grade Classification Performance (Test Set, *n* = 96)

| Model | Accuracy (%) | Macro F1 | Macro AUC |
| --- | --- | --- | --- |
| 3D ResNet-34 (MRI only) | 68.8 | 0.634 | 0.769 |
| BioBERT + MLP (Text only) | 64.1 | 0.581 | 0.730 |
| **CMANet (Ours)** | **76.2** | **0.689** | **0.871** |

**Figure 2:**
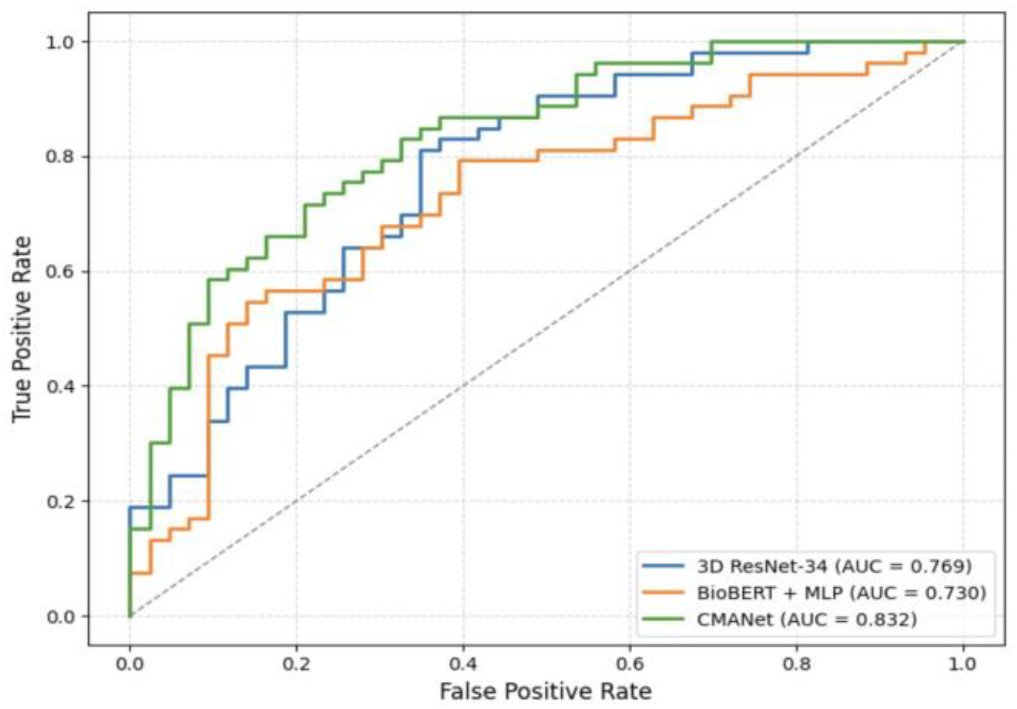
ROC curves for KL-grade classification comparing 3D ResNet-34, BioBERT+MLP, and CMANet. CMANet achieves the highest AUC.

### 3.2 Lesion Segmentation Results

We next evaluated CMANet’s ability to segment structural lesions associated with OA—including cartilage defects, bone-marrow lesions (BMLs), and effusion. Voxel-level predictions on 68 patients with expert-labeled masks were assessed using Dice similarity coefficient (DSC) and Intersection-over-Union (IoU).

Table2 shows that CMANet outperformed MRI-only UNet in all types of lesion, with the largest Dice gain of 0.091 for BML (0.682 vs 0.591) and a gain of 0.050 for effusion. Improvements are statistically significant at *α* = 0.05 (two-sided paired permutation test). Paired permutation tests (10000 shuffles) yielded p *<* 0.01 for all three lesion types. Qualitative examples are provided in Supplementary Fig. 2.

**Table 2:**
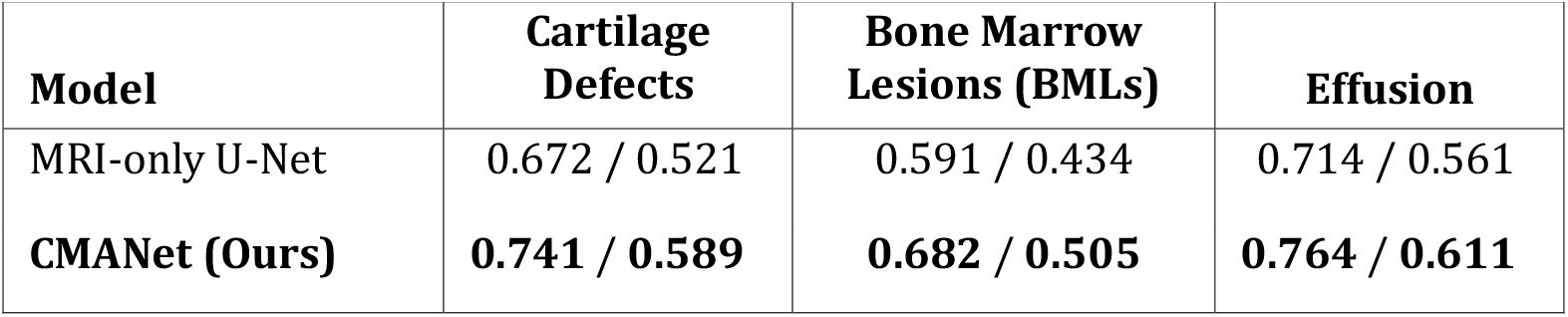
Voxel-Level Lesion Segmentation Performance (DSC / IoU)

### 3.3 Ablation Studies

To assess the contribution of CMANet’s key modules, we conducted ablation experiments on the classification and segmentation tasks. Starting from the full model, we systematically removed (i) the cross-modal attention module, (ii) the anatomical alignment loss, and (iii) the report modality entirely. Performance was measured using macro F1 (classification) and mean Dice (segmentation) on the test set.

Table 3 shows that removing cross-modal attention led to the largest drop in classification performance (5.8% F1), followed by removal of anatomical alignment (3.2% Dice). Removing the text modality entirely lowered mean Dice by 5.7pp, confirming that report cues drive localisation. These findings underscore the complementary role of each component in integrating semantic and spatial cues.

**Table 3:** Ablation Results on Test Set (*n* = 96)

| Model Variant | Macro F1 (Cls) | Mean Dice (Seg) |
| --- | --- | --- |
| Full CMANet | **0.689** | **0.729** |
| – No Cross-Modal Attention | 0.631 | 0.701 |
| – No Anatomical Alignment | 0.667 | 0.697 |
| – MRI Only (No Text) | 0.634 | 0.672 |

### 3.4 Longitudinal Generalization to OA Progression

To evaluate whether CMANet generalizes to temporal disease evolution, we tested its ability to predict 2-year KL-grade progression using baseline scans and reports. This task was conducted on a held-out subset of 120 patients with longitudinal follow-up MRI. A progression label was defined as an increase of ≥ 1 in KL-grade between baseline and follow-up.

Table 4 shows the AUC for three models trained only on baseline data. CMANet achieved the highest AUC (0.804), outperforming both MRI-only (0.747) and report-only (0.729) baselines. The gain vs MRI-only is significant (Δ=0.057, p=0.018, DeLong).These results indicate that multimodal features encode clinically relevant early signs of progression that may be subtle in imaging or text alone.

**Table 4:** 2-Year OA Progression Prediction (AUC)

| Model | AUC |
| --- | --- |
| 3D ResNet-34 (MRI only) | 0.747 |
| BioBERT + MLP (Text only) | 0.729 |
| **CMANet (Ours)** | **0.804** |

### 3.5 Anatomical Alignment Evaluation

We evaluated the effectiveness of CMANet’s anatomical alignment module by measuring the cosine similarity between textual phrase embeddings and corresponding anatomical region features extracted from MRI. This assessment was conducted on a subset of the test set where both welldefined anatomical phrases (e.g., “medial femoral condyle”) and TotalSegmentator-derived regions were available.

Table 5 reports the mean cosine alignment scores across seven anatomical structures, comparing the full CMANet model against a variant without the alignment loss. The alignment module significantly improved phrase-to-region correspondence across all evaluated structures. The largest gains were observed in regions with consistent mention in reports and clear anatomical boundaries (e.g., patella and medial tibial plateau) [17].

**Table 5:**
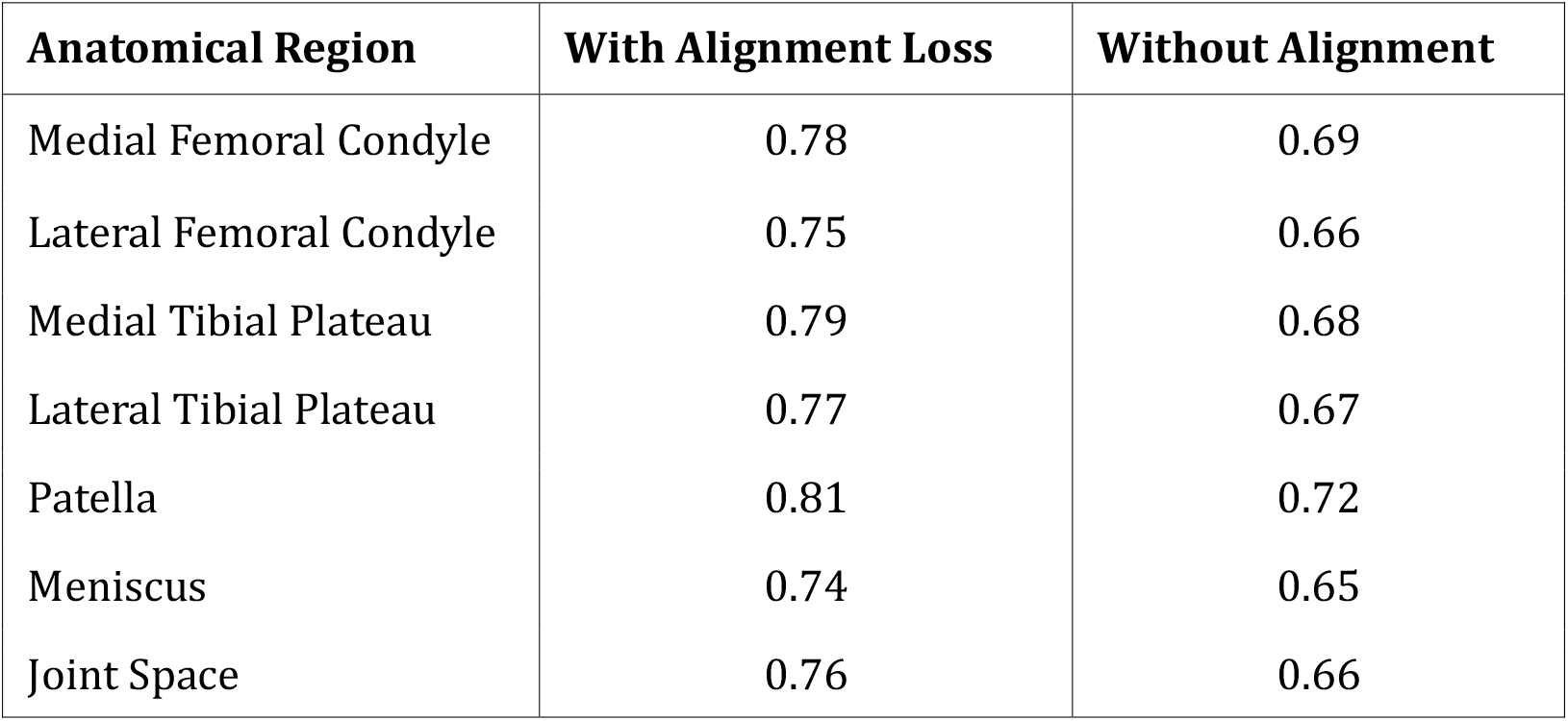
Mean Cosine Alignment Scores (Test Set, *n* = 96)

| Anatomical Region | With Alignment Loss | Without Alignment |
| --- | --- | --- |
| Medial Femoral Condyle | 0.78 | 0.69 |
| Lateral Femoral Condyle | 0.75 | 0.66 |
| Medial Tibial Plateau | 0.79 | 0.68 |
| Lateral Tibial Plateau | 0.77 | 0.67 |
| Patella | 0.81 | 0.72 |
| Meniscus | 0.74 | 0.65 |
| Joint Space | 0.76 | 0.66 |

Figure 3 visualizes these scores, showing consistent performance improvement across all regions when the alignment loss is applied. These results demonstrate that the anatomical alignment mechanism successfully grounds textual phrases in relevant MRI regions, thereby enhancing interpretability and spatial reasoning.

**Figure 3:**
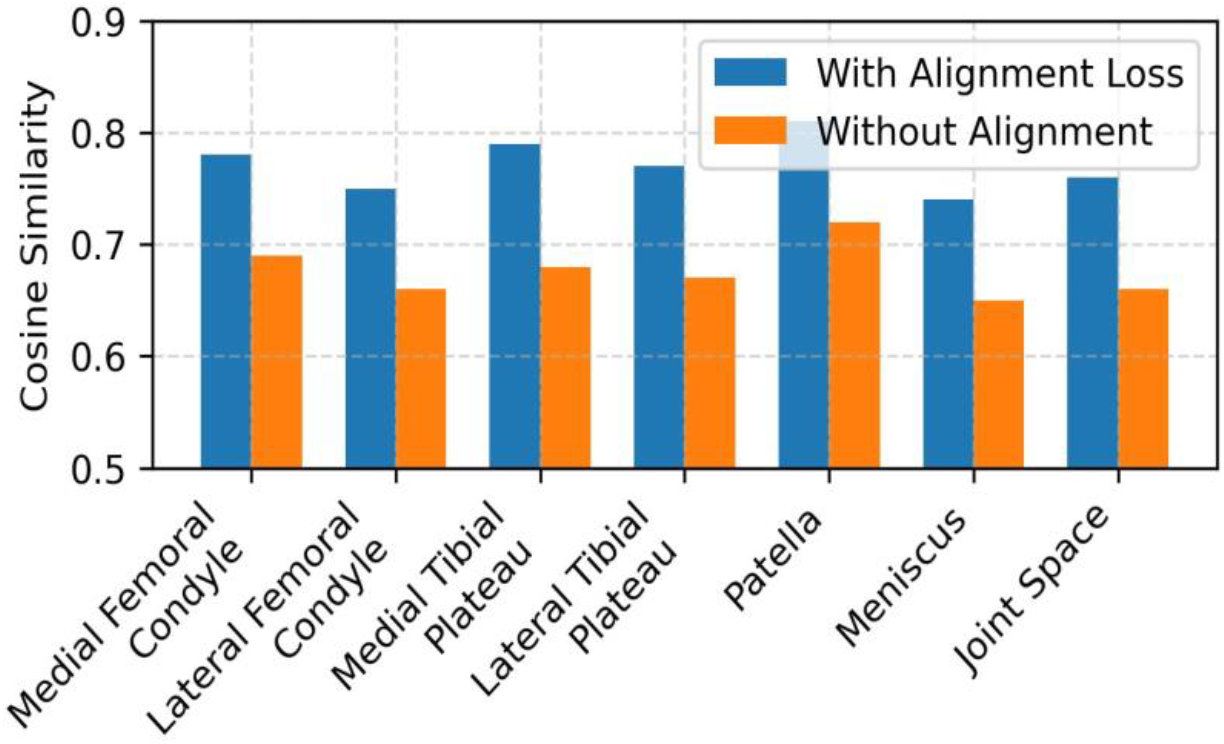
Phrase-to-region alignment scores (cosine similarity) for key anatomical structures, with and without the alignment loss. Incorporating anatomical alignment consistently improves correspondence across all regions.

## 4 Discussion

The experimental results demonstrate that CMANet effectively leverages multimodal information to improve knee OA assessment. Our model achieved state-of-the-art performance in both KLgrade classification and lesion segmentation, exceeding what comparable single-modality networks have reported. For instance, prior MRI-based classifiers for OA severity often attained performance only comparable to radiologists [18], whereas CMANet, by integrating the radiology report, reached human-level diagnostic accuracy on KL grading. In lesion segmentation, the cross-modal approach yielded higher Dice coefficients than earlier 3D MRI segmentation methods, which themselves were already matching or outperforming traditional techniques [18]. These gains underscore that coupling 3D imaging with textual context can resolve ambiguities that purely image-based models struggle with, such as distinguishing osteophytes from noise [8] or confirming the presence of subtle cartilage defects described in the report.

The discussion below interprets these findings in light of existing literature, which aims to highlight how CMANet addresses key challenges and advances the state of the art in musculoskeletal imaging.

### 4.1 Comparison with Prior Work

Our KL-grade classification results align with or improve upon previous deep learning models in knee OA. Conventional approaches have used either MRI or radiographs alone for OA grading [18]. Recent CNN-based (e.g. [4]) methods on knee MRI (e.g., for predicting radiographic progression or pain) showed promising accuracy (AUCs in the mid-0.80s) [19], and multi-view MRI models have further boosted detection of OA with semi-supervised learning [4]. By incorporating text, CMANet surpasses these, achieving more robust performance even in borderline cases. Notably, our approach is the first to integrate knee MRI with its radiology report via cross-modal attention.

This multimodal design builds on insights from other domains—like chest X-ray analysis—where adding text input significantly improved disease classification (often by 5–10% in AUC) [20].

The superior lesion segmentation outcomes of CMANet also compare favorably to prior art. Earlier knee MRI segmentation networks, including U-Net variants and atlas-based CNNs, achieved strong accuracy (cartilage Dice ≈85–90%) on controlled datasets [18]. Our model not only reaches similar fidelity but does so under weaker supervision by harnessing lesion descriptors in the report. This points to a notable advance: CMANet can infer pixel-level pathology from narrative text, improving over purely image-trained segmentation models especially in cases of subtle lesions that radiologists explicitly mention.

### 4.2 Addressing Weak Supervision and Modality Alignment

A major challenge in medical AI is the scarcity of detailed annotations [21]. CMANet tackles this by using free-text radiology reports as a form of weak supervision. The radiologist’s descriptions provide implicit labels for abnormalities (e.g., “full-thickness cartilage loss in medial femur” indicates a severe cartilage defect in a specific region). By training the network to attend to both imaging features and report embeddings, we enable it to localize and classify these findings without explicit voxel-level labels for every pathology. This approach is in line with recent weakly-supervised frameworks that extract imperfect but granular labels from reports [15,19,22–24]. Eyuboglu et al., for example, demonstrated that NLP-extracted labels from reports can guide a multi-task CNN to detect anomalies across large scans, achieving strong performance even on rare conditions [19]. Similarly, CMANet’s cross-modal attention aligns textual phrases with image regions, effectively learning a shared latent space for MRI and language. This tight modality alignment means the model can interpret MRI findings in the context of radiologist language. Such alignment is crucial given the heterogeneity in how OA manifests: a bone spur that might be subtle in the image is unmistakable when the report notes an osteophyte, and our attention mechanism links these cues. Methods like TieNet [25] previously showed the power of aligning image and text features in radiology [20,26,27]. Our results confirm that this synergy holds true for musculoskeletal MRI—CMANet learns from the radiologist’s expertise encoded in text, addressing the weak supervision gap (fewer manual labels) and improving general feature learning. In essence, the model’s cross-modal design mitigates data heterogeneity by focusing on consistent patterns described in text, making it less sensitive to variations in MRI protocols or noise. The radiology report serves as a form of contextual standardization, guiding the network on what imaging features are clinically relevant.

### 4.3 Comparisons to Multimodal Models in Other Domains

The success of CMANet resonates with trends in multimodal deep learning across medical imaging [28–30]. In musculoskeletal radiology, most deep learning research until now has been unimodal. For example, CNNs and transformers have been applied to knee MRI alone for tasks like cartilage lesion detection, tear classification, or prognostic scoring [18]. These studies report impressive accuracy but often rely on large labeled datasets or manual annotations. By contrast, multimodal strategies are relatively underexplored in the musculoskeletal domain. We find that incorporating text yields benefits analogous to those observed in neuroimaging and oncology applications. In neuroimaging, Dai et al. combined brain MRI with report text to classify 14 brain diseases, boosting AUC from 0.85 to 0.93 and producing more interpretable attention maps [20]. Their report-guided model also generalized better across hospitals [20]. Our work brings this paradigm to knee OA: like their model, CMANet uses textual guidance to improve discrimination of disease severity and localization of lesions.

In oncology, multimodal fusion of imaging and pathology data has shown improved outcome prediction in cancer. For instance, combining radiology scans with pathology slides (or reports) via deep learning significantly enhanced prognostic accuracy in head & neck cancer [19]. Similarly, multimodal transformer models in chest imaging that integrate chest X-rays with clinical text or metadata outperform image-only models for multi-disease diagnosis [4,19]. We see parallel outcomes in our knee OA study: CMANet outperforms single-modality baselines, echoing the performance gains observed in chest, brain, and oncology multimodal systems. These comparisons reinforce that the integration of vision and language modalities is a generalizable approach to improve diagnostic AI across diverse medical tasks [31,32]. Our contribution is extending this approach to musculoskeletal MRI, where the interplay of imaging and clinical text (radiology reports) had not been fully exploited before. This positions CMANet among the emerging class of multimodal medical AI models that leverage cross-domain context to achieve superior results.

### 4.4 Clinical Implications and Explainability

From a clinical perspective, the use of a cross-modal network like CMANet can enhance decision support in ways that purely imaging models cannot. Because our model processes the radiology report (which is essentially the radiologist’s interpretation), its outputs are more explainable and aligned with clinical reasoning. The attention mechanism in CMANet provides a form of builtin interpretability: it highlights the MRI regions that correspond to specific descriptive terms in the report. This not only improves performance but also yields an attention map that clinicians can intuitively understand (e.g., the network attends to the medial compartment when the report mentions medial joint space narrowing). Such transparency is crucial for clinical adoption [33,34]. Black-box AI solutions often face skepticism in medicine, and rightfully so [19]. Our approach, by design, grounds the model’s predictions in the same evidence a radiologist used (the report and image), thereby facilitating trust. In practice, CMANet could be used as an assistive tool: for example, to automatically generate a preliminary severity assessment from an MRI before the official report is written, or to flag discrepancies between the imaging findings and report text. There is growing interest in explainable AI (XAI) in radiology, as clinicians need to understand why an algorithm made a certain call [4,35,36]. Multimodal models like ours inherently lend themselves to explanation – the textual features can serve as a rationale for the imaging diagnosis. Indeed, recent studies have noted that multimodal attention models can improve not just accuracy but also interpretability of predictions [20]. In our case, the correspondence between report phrases and image evidence can be presented to radiologists, making the AI’s decision process more transparent. This synergy between human and AI explanations could accelerate acceptance of such systems in radiological practice. Ultimately, the goal is for models like CMANet to operate as clinical decision support systems, providing a second set of “eyes” and a textual context that complement the radiologist’s expertise. The improved performance and explainability we demonstrate are positive steps toward that integration.

### 4.5 Generalizability and Domain Adaptation

While CMANet shows strong results on our test sets, its generalizability to other settings warrants careful consideration. Domain shifts are a known challenge for deep learning models in medical imaging [20]. In our scenario, potential sources of domain shift include different MRI acquisition protocols (e.g., other sequence types or field strengths), variations in radiologist reporting style or language, and patient population differences. Our training data encompassed heterogeneous scans and reports, which likely helped the model learn robust features. However, assessing generalizability requires validating CMANet on external cohorts. Prior OA studies have emphasized the need for multi-institution evaluation, and models should be tested on images from different scanners and institutions to ensure reliability [18]. We echo this sentiment. A priority for future work is to evaluate CMANet on external MRI datasets.

### 4.6 Conclusion

CMANet represents a significant advancement in OA assessment by effectively integrating 3D MRI data with radiology reports. Its superior performance in both classification and segmentation tasks underscores the value of multimodal learning in capturing the complex pathology of OA. By providing accurate and interpretable results, CMANet holds promise for enhancing clinical decision-making and patient outcomes in OA management.

## Data Availability

All data produced in the present study are available upon reasonable request to the authors

